# Comprehensive Analysis of Therapeutic Strategies Using Mesenchymal Stem Cell-Derived Exosomes in Preclinical Models of Osteoarthritis

**DOI:** 10.1101/2024.09.19.24313971

**Authors:** Riya Mukherjee, Ramendra Pati Pandey, Gunjan, Himanshu, Ing-Kae Wang, Sing-Ying Hsieh, Chung-Ming Chang

**Affiliations:** Graduate Institute of Biomedical Sciences, Chang Gung University; School of Health Sciences and Technology (SoHST), UPES Dehradun, Uttarakhand, India; Cell Culture Medium Technology Development, Regeneration Medicine Technology Division, Biomedical Technology and Device Research Laboratories, ITRI, Chutung, Hsinchu, Taiwan 310401, R.O.C; Master & Ph.D Program in Biotechnology Industry, Chang Gung University; Department of Medical Biotechnology and Laboratory Science; Chang Gung University

**Keywords:** Osteoarthritis, Exosome Therapy, Mesenchymal Stem Cells, Preclinical Studies, Therapeutic Efficacy

## Abstract

**Background:** Osteoarthritis (OA) is a common degenerative disease affecting people and animals, resulting in persistent pain and joint deformities. Its growing prevalence presents considerable difficulties to public health and veterinary care systems worldwide. Despite substantial research, the molecular pathways underlying OA pathogenesis remain poorly understood, limiting the development of effective treatment strategies. Exosomes, or small endosomal membrane microvesicles, have emerged as intriguing vehicles for intercellular communication and medicinal administration in a variety of illnesses, including OA. However, their efficacy and action methods in preclinical OA models require additional exploration.

**Methods:** We analyzed several databases from 2016 to 2023 for original studies on exosome treatment in preclinical OA models. The inclusion criteria included studies that used exosomes generated from mesenchymal stem cells (MSCs) in both human and animal models of OA. Thematic synthesis and data extraction were used to examine research features, dosage administration techniques, and efficacy results. The quality of included studies was assessed using recognized criteria, and statistical analysis was performed to determine the efficacy of exosome treatment in decreasing Osteoarthritis Research Society International (OARSI) scores.

**Results:** Our study comprised thirteen peer-reviewed articles that included both human and animal models of OA. Most trials used bone marrow MSC-derived exosomes administered intra-articularly. The analysis of OARSI scores revealed a considerable reduction in joint deterioration following exosome therapy. Source analysis demonstrated that exosome treatment originating from human and animal MSCs was consistently effective. However, an assessment of study quality revealed potential biases and limitations, emphasizing the need for more research to validate these findings and refine therapy options for OA management.

Graphical Abstract

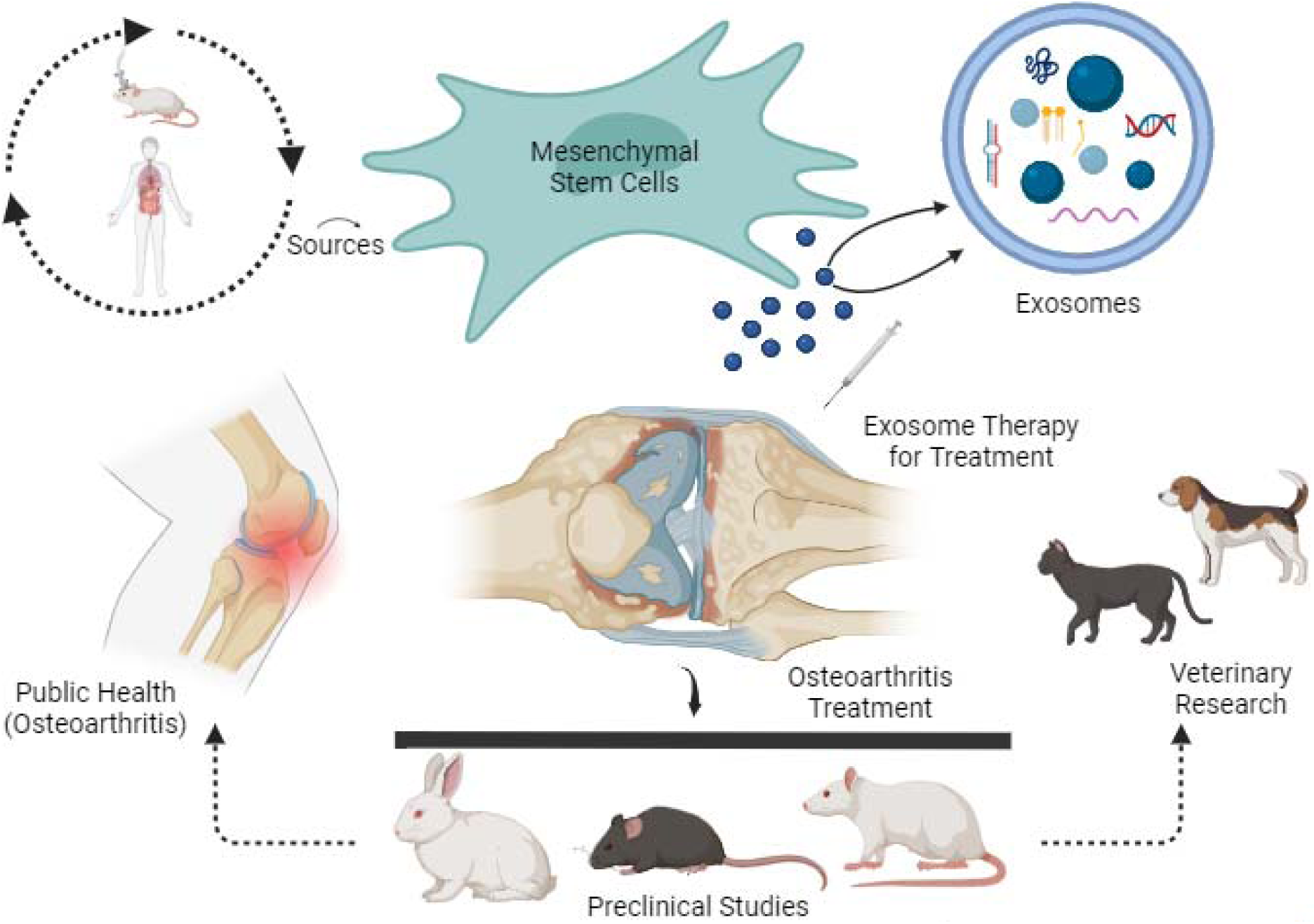

## 1. Introduction

Osteoarthritis (OA) is the most common degenerative disease both in humans and in animals, a leading cause of disability, and is characterized by chronic pain and a range of joint abnormalities, including articular cartilage damage, synovial inflammation, subchondral bone remodeling, and the formation of osteophytes (Steinmetz et al., 2023; Tong et al., 2022). The incidence of OA is steadily increasing due to the rising aging population and the global obesity epidemic, imposing a significant socioeconomic burden and posing a significant challenge to public health (A. R. Sun et al., 2021; Wen & Xiao, 2022; X. Wu et al., 2022). The World Health Organization (WHO) has designated the period from 2021 to 2030 as the "Decade of Healthy Ageing," with a particular emphasis on enhancing both life expectancy and overall quality of life. This designation presents an important opportunity to address the burden of OA within the framework of adult health (WHO). Due to the chronic nature of osteoarthritis and its substantial impact on mobility and daily functioning, this is especially important. According to a WHO report, in 2019, approximately 528 million people worldwide were living with OA, marking a 113% increase since 1990. Most individuals affected by OA, about 73%, were aged 55 and older, with females accounting for 60% of the total. With 365 million incidents, the knee emerged as the most affected joint, followed by the hip and the hand (WHO, 2023). A population-based meta-analysis published in a Chinese publication in 2019 combined existing papers to determine the prevalence of knee joint osteoarthritis (OA) in China. The investigation found that the prevalence of knee joint OA in China stood at 17%, with a breakdown of 12.3% in men and 22.2% in women (X. Sun et al., 2019). In a separate study conducted in 2019, which compiled and analyzed existing publications, the research identified lumbar joint OA as the most widespread type of OA. It exhibited a prevalence rate of 25.03% in Chinese middle-aged and elderly populations. Knee joint OA came in second with a prevalence of 21.51%, followed by cervical joint OA at 20.46%, and hand joint OA at 8.99% (X. Sun et al., 2019). Additionally, in Taiwan, knee OA has a prevalence rate of approximately 15%, impacting more than 3.5 million individuals who experience joint pain (DuoGenicStemCellscorporation, 2023). Furthermore, In India, from 1990, OA, number rose from approximately 23.46 million people to 62.35 million by 2019 (Singh et al., 2022). According to the CDC, 32.5 million US individuals have OA, and 1 in 4 (or 54.4 million) US adults have some form of arthritis. By the year 2040, 78 million people will be predicted to have arthritis worldwide (Alliance). In Western Europe, OA affected approximately 57 million people in 2019 and cost over 2 million years of healthy life. Since 1990, the number of people affected in the area has increased by 54% (Economist, 2019). Besides, in US, 63.4 million households have a dog, and there are an estimated 9 million pet dogs owned there (PFMA, 2018), Osteoarthritis affects many dogs globally and is a danger to the wellbeing of dogs. According to information gathered from 200 veterinarians, 20% of all dogs in North America over the age of one are estimated to have osteoarthritis (SA, 1997). Recent estimates suggest that osteoarthritis affects 2.5% to 6.6% of dogs of all ages and breeds visiting primary care offices in the United Kingdom (Pye et al., 2024).Canine osteoarthritis is a significant concern for veterinarians, owners, and breeders worldwide, and it has a considerable impact on the welfare of dogs. The financial burden of treatment options for canine osteoarthritis can be particularly challenging for dog owners.

At the molecular level, the pathogenesis of OA involves complex interactions between various cellular and molecular processes. These include dysregulation of chondrocyte metabolism, abnormal remodeling of the extracellular matrix, activation of inflammatory pathways, and disruption of homeostatic mechanisms. Despite decades of research, the exact causes and molecular mechanisms underlying OA are not fully understood, which contributes to the limited effectiveness of traditional therapeutic approaches in stopping the progression of the disease (Tong et al., n.d.). Chondrocytes, the cells responsible for maintaining cartilage integrity, play a central role in the pathogenesis of OA. Dysregulated chondrocyte metabolism leads to imbalances in extracellular matrix (ECM) turnover, characterized by decreased synthesis of collagen and proteoglycans, and increased expression of matrix metalloproteinases (MMPs) and aggrecanases. These enzymes degrade the ECM, compromising cartilage structure and function. Synovial inflammation is a defining feature of osteoarthritis and contributes to its development. OA joints exhibit higher levels of inflammatory mediators, including IL-1β, TNF-α, and PGE2. These cytokines enhance chondrocyte catabolism, accelerate cartilage breakdown, and drive synovial cell production of inflammatory mediators. In OA, dysregulated signaling pathways such NF-κB, MAPK, and Wnt/β-catenin contribute to inflammation, apoptosis, and ECM degradation. The activation of these pathways results in the expression of pro-inflammatory cytokines, catabolic enzymes, and cartilage breakdown mediators (Yao et al., n.d.).

Exosomes are small endosomal membrane microvesicles that have gotten a lot of interest in the last decade. Exosomes were discovered in extracellular space as early as the late 1980s (Y. Zhang, Liu, Liu, & Tang, 2019). Exosomes released by cells, on the other hand, were previously proposed as cellular waste resulting from cell injury or by-products of cell homeostasis, with no substantial impact on nearby cells (Y. Zhang et al., 2019). It was only recently that these extracellular vesicles were shown to be functional vehicles capable of conveying a complex cargo of proteins, lipids, and nucleic acids to the target cells they encountered, which may eventually reprogram the destination cells distal from their release (S.-p. Li, Lin, Jiang, & Yu, 2018). Thus, exosomes constitute a novel route of intercellular communication that may be important in a variety of cellular activities, including immunological response (Greening, Gopal, Xu, Simpson, & Chen, 2015), signal transduction (Gangoda, Boukouris, Liem, Kalra, & Mathivanan, 2015), and antigen presentation (Smith, Cheng, Bryant, & Schorey, 2017). Exosomes can be discharged by almost all eukaryotic cells, and their cargos are thought to differ substantially depending on the origin of the cell type and its current condition (e.g. transformed, differentiated, stimulated, and stressed). Thus, exosomes and their biologically active cargos may provide preemptive information in a variety of disorders, including neurodegenerative, and degenerative diseases (Howitt & Hill, 2016; W. Liu et al., 2019). Emerging studies demonstrated that, exosomes originating from mesenchymal stem cells (MSCs) provide protective effects for bone and cartilage in OA (Zhu et al., 2017). They achieve this by elevating the expression of chondrocyte markers such as type II collagen and aggrecan, suppressing inflammatory markers like nitric oxide synthase (iNOS), preventing chondrocyte apoptosis, and promoting both the migration and proliferation of chondrocytes (Zhu et al., 2017) and inhibiting the activation of macrophages (Cosenza, Ruiz, Toupet, Jorgensen, & Noël, 2017). Consequently, exosomes are believed to have a role in the development and progression of various diseases, including OA.

In the field of clinical practice, systematic reviews and meta-analyses, conducted with strict adherence to scientific standards, are considered the most objective and comprehensive methods for evaluating intervention effectiveness. To ensure the validity of our findings, we conducted a thorough analysis of the efficacy and likely mechanisms of exosome utilization in both human and animal pre-clinical models of osteoarthritis. Our study focused uniquely on osteoarthritis among the various degenerative diseases. We believe that this analysis of preclinical data will significantly contribute to shaping the design of future clinical trials. The efficacy of exosomes in preclinical settings presents intriguing opportunities for future research in the veterinary and public health fields.

## 2. Materials and methods

### 2.1. Data Collection

An extensive literature search was conducted from 2016 to 2023 by utilizing various databases, including Web of Science, Google Scholar, ScienceDirect, PubMed, ProQuest, EMBASE, and SCOPUS. The aim was to carry out a comprehensive analysis focused on the topics of ‘extracellular vesicles’ or ‘exosomes’ and their relevance to ‘osteoarthritis’. To construct our search strategy, we employed a combination of Medical Subject Headings (MeSH) terms, free-text keywords, and Boolean operators (AND or OR). The search strategy can be outlined as follows with the following keywords: (exosome OR microvesicles OR exosomes OR extracellular vesicles) AND (human OR (animal OR mouse OR rat OR rabbit)) AND (arthritis or osteoarthritis OR degenerative joint disease OR Degenerative arthritis OR degenerative joint disease OR Knee osteoarthritis). This search strategy was applied across different Medline databases. The design and reporting of this study adhered to the guidelines outlined in the Preferred Reporting Items for Systematic Reviews and Meta-Analysis (PRISMA) statement (Liberati, Altman, Tetzlaff, & Mulrow, 2009; A. Liberati et al., 2009).

### 2.2. Inclusion and exclusion criteria

Based on the specified inclusion and exclusion criteria below, a total of 13 papers were selected for retrieving information. Studies were included in the analysis if they met the following criteria: (i) original investigation in humans’ and animals’ pre-clinical models with osteoarthritis, (ii) exosome-based therapy, (iii) studies including author names, publication year, weight, number of dosages, dosage units, and route of administration. Articles undergoing full-text were excluded for the following reasons: (i) being review articles, (ii) inability to extract or obtain data from the original authors, (iii) incomplete data or data expressed as ratios or percentages, and (iv) not being published as a full-text article in a journal.

### 2.3. Data extraction

Data from various osteoarthritis (OA) treatments using exosomes were systematically compiled into a spreadsheet using Excel® (Microsoft® Office Excel 2013) and subjected to preliminary testing before complete extraction. The extracted information included the author’s name, country of publication, year of publication (2017–2023), mesenchymal stem cell source, isolation method, exosome size, tissue origin, exosome immunocompatibility, and associated degenerative disease. Additionally, another spreadsheet was created to include author’s name, dosage administration, route of application, number of dosages, age, weight, and OARSI scores. The collected data were analyzed further, and details related to OA treatment strategies were meticulously organized into tables. Citations from the compiled papers were managed using Endnote 20 (Clarivate Analytics, U.S.A).

### 2.4. Quality assessment

Assessment of the reporting quality of included studies was performed using a scoring system adapted from (Papazova et al., 2015; Wever et al., 2012). Evaluating publication bias is essential to ensure the integrity and validity of our meta-analysis in assessing the effects of exosomes on osteoarthritis (OA). Our approach aimed to systematically address any potential bias, ensuring that our meta-analysis offers an impartial synthesis of the existing evidence on the therapeutic benefits of exosomes in OA. Furthermore, we employed a risk bias checklist across multiple domains (D1 to D5), with each domain evaluated using five categories: high risk, some concerns, low risk, no information, and not applicable. Evaluation of the risk of bias was performed using version 2 of the Cochrane risk-of-bias tool for randomized trials (RoB 2) (Sterne JAC, 2019).

### 2.5. Statistical analysis

The statistical analysis and generation of forest plots and heat maps for pooled summary estimates were conducted using the meta or metaphor and stats package in R software. The summary estimates were derived from pooled data of forest plots representing Osteoarthritis Research Society International (OARSI) scores in the OA model of humans and animals before and after exosome treatment. 95% confidence intervals (CI) and odds ratios (OR) for both fixed-effect and random-effects models were calculated. OARSI is a widely used indicator of success in research on osteoarthritis. It was created by international specialists in the field working in consensus, and it has been approved for use in clinical trials. Making use of a standardized outcome measure, such as the OARSI, can assist assure consistency between research and enable useful comparisons. Utilizing OARSI can also make it easier to combine data from several research, improving the meta-analysis’s statistical power and accuracy. The I^2^ and τ^2^ statistics were used to evaluate the heterogeneity of the data. Random-effect models were employed for all meta-analyses, and the outcomes were presented in forest plots. Additionally, the statistical significance was determined by p-value (p ≤ 0.05).

## 3. Results

### 3.1. Study Selection

A total of 1280 papers published between 2016 and 2023 were systematically categorized using Medline databases i.e. PubMed, Google Scholar, Scopus, and ISI Web of Science. Following a preliminary assessment of titles and abstracts, 326 studies underwent initial screening. From these, 247 records were excluded due to irrelevance or redundancy, leaving 79 articles for further examination. Subsequently, 59 articles were excluded for various reasons: 10 articles were excluded for improper screening, 9 articles due to no relevant animal intervention, 32 articles were review articles, 2 were case studies, 2 were book chapters, and 4 articles could not be accessed Further proper screening process include 20 studies. Among these, 7 articles were excluded for the following reasons: in 5 articles, exosomes were not derived from mesenchymal stem cells, and 2 articles had incomplete data. Finally, 13 studies (Cosenza et al., 2017; He et al., 2020; Jin et al., 2021; X. Li et al., 2021; Liang et al., 2022; Lin et al., 2021; Y. Liu et al., 2022; Tao et al., 2017; Wang et al., 2021; J. Wu et al., 2019; J. Zhang, Rong, Luo, & Cui, 2020; Zhu et al., 2017). were selected for this meta-analysis. The study selection process adhered to the PRISMA flow diagram (A. Liberati et al., 2009), depicted in Figure 1.

**Figure 1.**
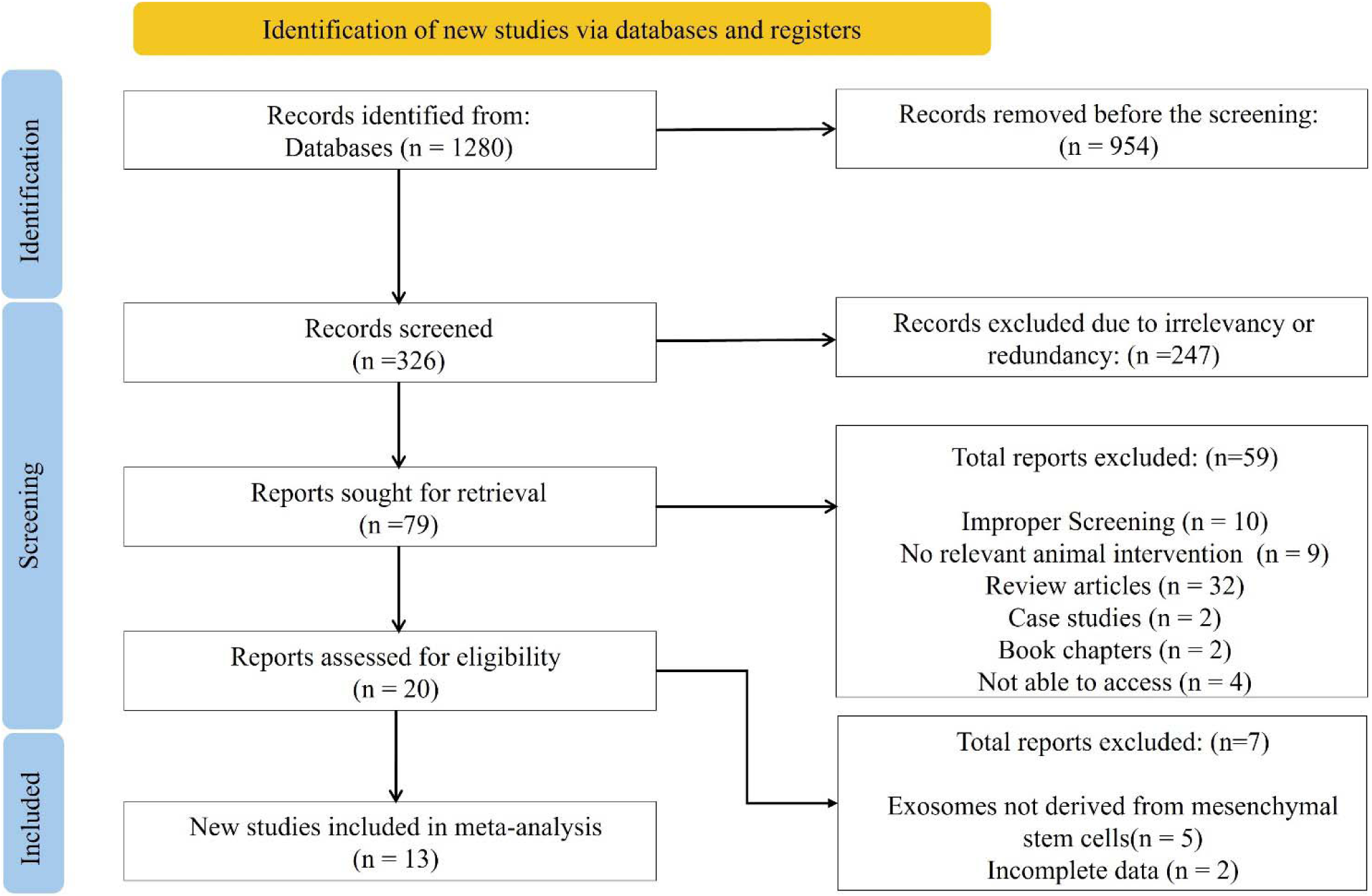
PRISMA chart depicting the inclusion criteria for studies investigating exosome therapy in preclinical models of osteoarthritis.

### 3.2. Study Characteristics

The study summarizes data from various studies on the use of exosomes derived from human mesenchymal stem cells (MSCs) for osteoarthritis (OA) treatment, spanning from 2017 to 2022 (**Table 1**). Most studies are from China and focus on human-derived MSCs. A common aspect among the studies is the use of ultracentrifugation as the primary method for exosome isolation, seen in the works of Zhu et al. (2017), Lin et al. (2020), Jin et al. (2021), Liu et al. (2022), Wang et al. (2021), Tao et al. (2017), and Li et al. (2022). Other methods include tangential flow filtration (Zhang et al., 2019) and the exoEasy Kit (Yang et al., 2022). The exosome sizes reported range from 30 to 200 nm. The sources of MSCs vary, including induced pluripotent stem cells (ipMSC), synovial membrane MSCs (SMMSC), dental pulp stem cells (DpSCs), bone marrow (BM), knee articular cartilage, human synovial tissue, embryonic stem cells, and umbilical cord (UC) MSCs.

**Table 1.**
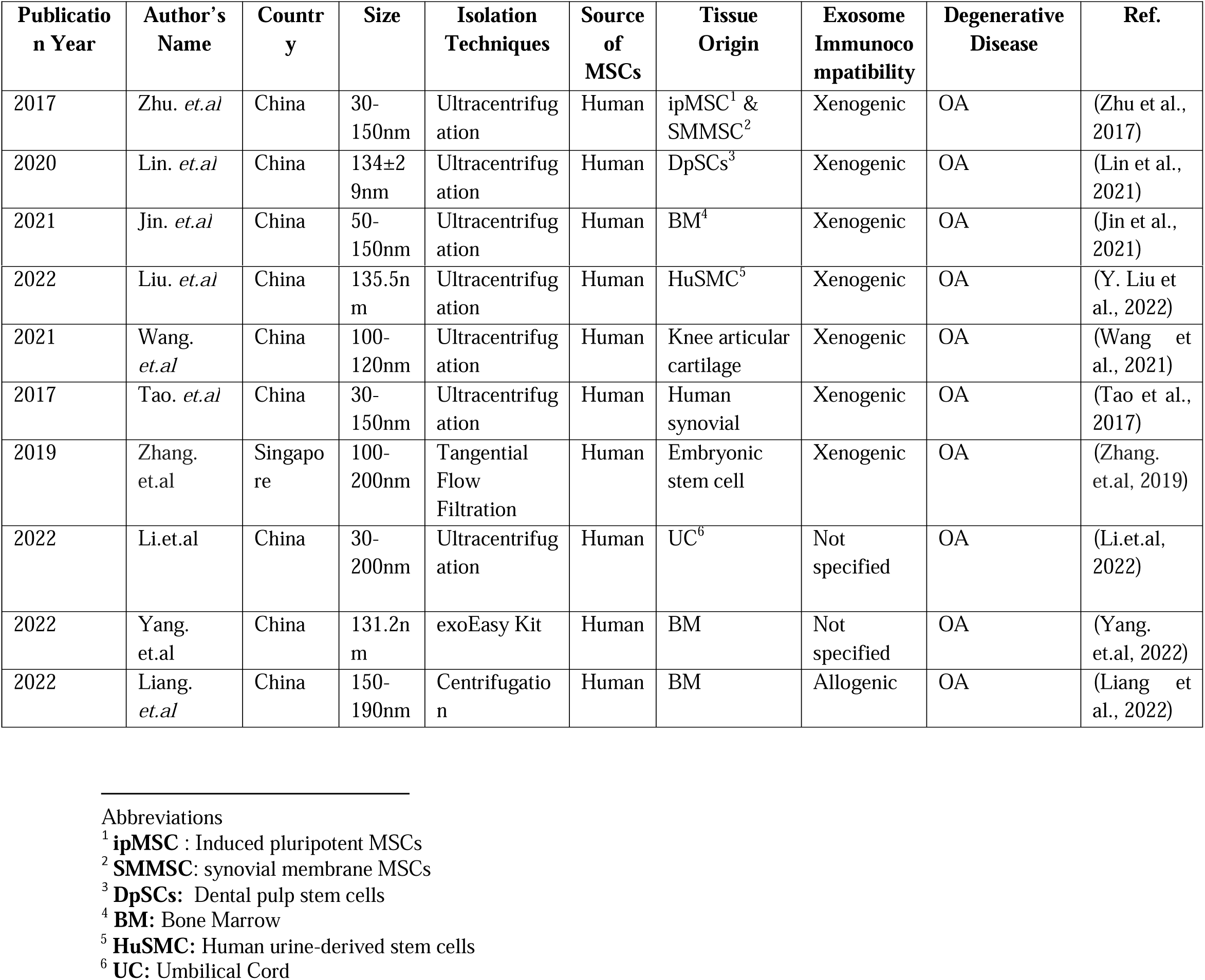
Interventional data of exosomes derived from human sources.

| Publication Year | Author's Name | Country | Size | Isolation Techniques | Source of MSCs | Tissue Origin | Exosome Immunocompatibility | Degenerative Disease | Ref. |
| --- | --- | --- | --- | --- | --- | --- | --- | --- | --- |
| 2017 | Zhu. *et.al* | China | 30-150nm | Ultracentrifugation | Human | ipMSC<sup>1</sup> & SMMSC<sup>2</sup> | Xenogenic | OA | (Zhu et al., 2017) |
| 2020 | Lin. *et.al* | China | 134±29nm | Ultracentrifugation | Human | DpSCs<sup>3</sup> | Xenogenic | OA | (Lin et al., 2021) |
| 2021 | Jin. *et.al* | China | 50-150nm | Ultracentrifugation | Human | BM<sup>4</sup> | Xenogenic | OA | (Jin et al., 2021) |
| 2022 | Liu. *et.al* | China | 135.5nm | Ultracentrifugation | Human | HuSMC<sup>5</sup> | Xenogenic | OA | (Y. Liu et al., 2022) |
| 2021 | Wang. *et.al* | China | 100-120nm | Ultracentrifugation | Human | Knee articular cartilage | Xenogenic | OA | (Wang et al., 2021) |
| 2017 | Tao. *et.al* | China | 30-150nm | Ultracentrifugation | Human | Human synovial | Xenogenic | OA | (Tao et al., 2017) |
| 2019 | Zhang. *et.al* | Singapore | 100-200nm | Tangential Flow Filtration | Human | Embryonic stem cell | Xenogenic | OA | (Zhang. et.al, 2019) |
| 2022 | Li.et.al | China | 30-200nm | Ultracentrifugation | Human | UC<sup>6</sup> | Not specified | OA | (Li.et.al, 2022) |
| 2022 | Yang. *et.al* | China | 131.2nm | exoEasy Kit | Human | BM | Not specified | OA | (Yang. et.al, 2022) |
| 2022 | Liang. *et.al* | China | 150-190nm | Centrifugation | Human | BM | Allogenic | OA | (Liang et al., 2022) |

In terms of immunocompatibility, most exosomes are characterized as xenogenic, except for Liang et al. (2022), who reported allogenic exosomes. The studies uniformly target OA as the degenerative disease of interest. Despite variations in exosome sizes and specific MSC sources, the overarching goal remains the same: exploring the therapeutic potential of MSC-derived exosomes in OA treatment. The detailed scrutiny and organization of these studies into tables enable a comprehensive understanding of OA treatment strategies using exosomes, as well as the methodologies and outcomes associated with each study.

Table 2, summarizes interventional data from studies on exosomes derived from animal sources for osteoarthritis (OA) treatment, spanning from 2017 to 2020 and involving researchers from France and China. Common across these studies is the use of centrifugation for exosome isolation, with Cosenza et al. (2017) and He et al. (2020) using standard centrifugation, and Zhang et al. (2020) employing ultracentrifugation. The MSCs were sourced from rodents, specifically mice (Cosenza et al., 2017) and rats (He et al., 2020; Zhang et al., 2020), with bone marrow (BM) being the primary tissue origin. Exosome sizes ranged from 96 nm to 153 nm, and all exosomes were allogenic, indicating derivation from the same species but different genetic backgrounds. Each study uniformly focused on OA, highlighting a shared interest in the therapeutic potential of animal-derived MSC exosomes for this condition. The table’s detailed organization facilitates a comprehensive understanding of OA treatment strategies using these exosomes.

**Table 2.** Interventional data of exosomes derived from animal sources.

| Publication Year | Author's Name | Country | Size (nm) | Isolation Techniques | Source of MSCs | Tissue Origin | Exosome Immunocompatibility | Degenerative Disease | Ref |
| --- | --- | --- | --- | --- | --- | --- | --- | --- | --- |
| 2017 | Cosenza. *et.al* | France | 96 | Centrifugation | Mouse | MBM<sup>7</sup> | Allogenic | Osteoarthritis | (Cosenza et al., 2017) |
| 2020 | He. *et.al* | China | 153 | Centrifugation | Rat | BM | Allogenic | Osteoarthritis | (He et al., 2020) |

<sup>5</sup> **HuSMC:** Human urine-derived stem cells
<sup>6</sup> **UC:** Umbilical Cord
<sup>7</sup> **MBM:** Murine Bone Marrow

|  |  |  |  |  |  |  |  |  |  |
| --- | --- | --- | --- | --- | --- | --- | --- | --- | --- |
| 2020 | Zhang.<br>*et.al* | China | 140 | Ultracentrifug<br>ation | Rat | BM | Allogenic | Osteoarthritis | (J. Zhang et<br>al., 2020) |

### 3.3. Dosage Administration

The studies investigating the therapeutic efficacy of mesenchymal stem cell-derived exosomes in preclinical models of osteoarthritis (OA). Each study is characterized by specific parameters, including dosage, administration route, number of dosages, age and weight of the animal models, and the Osteoarthritis Research Society International (OARSI) scores before and after exosome treatment (**Table 3**). Notably, the studies emphasize the use of exosomes derived from human sources, highlighting their potential therapeutic importance. Zhu et al. (2017) administered 8 μl of human-derived exosomes at a concentration of 1.0 × 10^10/ml via intra-articular injections, with two doses given to 6-week-old animals. The primary outcome showed a significant reduction in OARSI scores from 5 in the OA model to 1.75 after exosome treatment, indicating a substantial therapeutic effect. Lin et al. (2021) employed a single intra-articular injection of 100 μl human-derived exosomes in animals weighing approximately 358 ±5 grams, reporting a primary outcome of a decrease in OARSI scores from 2.3 in the OA model to 1.25 post-treatment. Jin et al. (2021) used 50 μl of human-derived exosomes at a concentration of 5 × 10^10/ml, administered via intra-articular injections, with two doses given to 8-week-old animals weighing 200±20 grams. The primary outcome recorded a reduction in OARSI scores from 5.8 in the OA model to 2 after exosome treatment. Liu et al. (2022) applied two intra-articular injections of 100 μl human-derived exosomes to 12-week-old animals weighing 250±20 grams. This research demonstrated a primary outcome of a significant decrease in OARSI scores from 20.5 in the OA model to 11.5 after treatment. Wang et al. (2021) conducted their study using 30 μl human-derived exosomes administered intra-articularly in two doses, though the age and weight of the animals were not specified. The OARSI scores were reduced from 21.2 in the OA model to 10.5 following treatment. Lastly, Tao et al. (2017) administered 100 μl of human-derived exosomes via intra-articular injections in two doses to 12-week-old animals weighing between 300-350 grams. This study observed a decrease in OARSI scores from 22 in the OA model to 9.25 after exosome treatment.

**Table 3.** Dosage administration of human-derived exosome treatment for OA.

| Author's<br>Name | Dosage Administration |  | Administration<br>Route | No. of<br>dosage | Age<br>(weeks) | Weight | OARSI Scores |  |  | Ref |
| --- | --- | --- | --- | --- | --- | --- | --- | --- | --- | --- |
|  | Control | Exosome<br>Treatment |  |  |  |  | Control | OA<br>Model | After<br>Exosome<br>Treatment |  |
| Zhu.<br>*et.al* | 8 μl | ($1.0 \times 10^{10}$/ml) | Intra-articular | 2 | 6 | NG | 0.3 | 5 | 1.75 | (Zhu et<br>al.,<br>2017) |
| Lin. *et.al* | 100 μl | 100 μl | Intra-articular | 1 | NG | $358 \pm 5$g | 0.25 | 2.3 | 1.25 | (Lin et<br>al., |
|  |  |  |  |  |  |  |  |  |  | 2021) |
| Jin. *et.al* | 50 µl | 5 x 10<sup>10</sup>/ml | Intra-articular | 2 | 8 | 200±20g | 4.3 | 5.8 | 2 | (Jin et al., 2021) |
| Liu. *et.al* |  | 100µl | Intra-articular | 2 | 12 | 250±20g | 0.25 | 20.5 | 11.5 | (Y. Liu et al., 2022) |
| Wang. *et.al* | 30µl | 30µl | Intra-articular | 2 | NG | NG | 0.25 | 21.2 | 10.5 | (Wang et al., 2021) |
| Tao. *et.al* | 100µl | 100µl | Intra-articular | 2 | 12 | 300-350g | 0.2 | 22 | 9.25 | (Tao et al., 2017) |

Comparatively, animal-derived exosomes also exhibited therapeutic efficacy but with varying degrees of success (**Table 4**). For instance, Cosenza et al. (2017) observed a reduction in OARSI scores from 11 to 5.8, and Liang et al. (2020) reported a decrease from 12.5 to 7.54. These findings underscore the consistent and effective results of exosome treatments in reducing OA severity, with human-derived exosomes showing a slight edge due to more consistently effective outcomes across multiple studies. This highlights the promising potential of both human and animal-derived exosomes, emphasizing the need for further research to fully elucidate the therapeutic mechanisms and optimize the clinical application of exosome-based therapies in OA treatment.

**Table 4.** Dosage administration of animal-derived exosome treatment for OA.

| Author's Name | Dosage units | Route of administration | No. of dosage | Age (weeks) | Weight | OARSI Scores |  |  | Ref |
| --- | --- | --- | --- | --- | --- | --- | --- | --- | --- |
|  |  |  |  |  |  | Control | OA Model | After Exosome Treatment |  |
| Cosenza. *et.al* | 200ng | Intra-articular | 3 | 10 | NG | 1.5 | 11 | 5.8 | (Cosenza et al., 2017) |
| Zhang. *et.al* | 40µl | Intra-articular | 1 | NG | 200-250g | 0.8 | 3.1 | 1.5 | (He et al., 2020) |
| He. *et.al* | 40µg | Intra-articular | 1 | 10 | NG | 1 | 4.1 | 3 | (Liang et al., 2022) |
| Liang. *et.al* | 50 or 100ug | Intra-articular | 1 | 6 | NG | 0.3 | 12.5 | 7.54 | (J. Zhang et al., 2020) |

### 3.3. Analysis of Exosome Therapy Derived from Human and Animal Sources

The finding presents a comprehensive analysis of the overall efficacy of exosome treatments on osteoarthritis (OA) across all included studies. This forest plot **Figure 2**, aggregates data from both human and animal-derived exosomes, providing a pooled estimate that reflects the general effectiveness of these therapies in managing OA. The pooled estimate indicates a significant positive effect of exosome therapy, with an odds ratio (OR) of 0.51 (95% CI: 0.41 to 0.61), and a p-value of less than 0.01, demonstrating the therapy’s effectiveness in reducing OA severity. However, substantial heterogeneity is observed among the studies (I² = 94%), suggesting considerable variability in effect sizes. In the studies for human-derived exosomes, Zhu et al. (2017) reported an OR of 0.34 (95% CI: 0.21 to 0.49), contributing 8.9% to the overall weight. Tao et al. (2017) observed an OR of 0.52 (95% CI: 0.31 to 0.73), with a weight of 7.2%. Lin et al. (2020) reported an OR of 0.50 (95% CI: 0.31 to 0.69), contributing 7.8% to the weight. Jin et al. (2021) showed an OR of 0.50 (95% CI: 0.43 to 0.56), with a weight contribution of 10.2%. Wang et al. (2021) observed an OR of 0.56 (95% CI: 0.49 to 0.63), contributing 10.2% to the weight. Liu et al. (2022) reported an OR of 0.42 (95% CI: 0.35 to 0.49), with a weight contribution of 10.2%. Li et al. (2022) showed the highest OR of 0.88 (95% CI: 0.80 to 0.93), contributing 10.3% to the weight. Similarly, in the studies for animal-derived exosomes, Cosenza et al. (2017) reported an OR of 0.38 (95% CI: 0.21 to 0.58), contributing 7.9% to the overall weight. He et al. (2020) showed an OR of 0.34 (95% CI: 0.22 to 0.48), with a weight contribution of 9.1%. Zhang et al. (2019) reported an OR of 0.50 (95% CI: 0.41 to 0.59), contributing 9.8% to the weight. Liang et al. (2022) observed an OR of 0.62 (95% CI: 0.46 to 0.77), contributing 8.5% to the weight.

**Figure 2.**
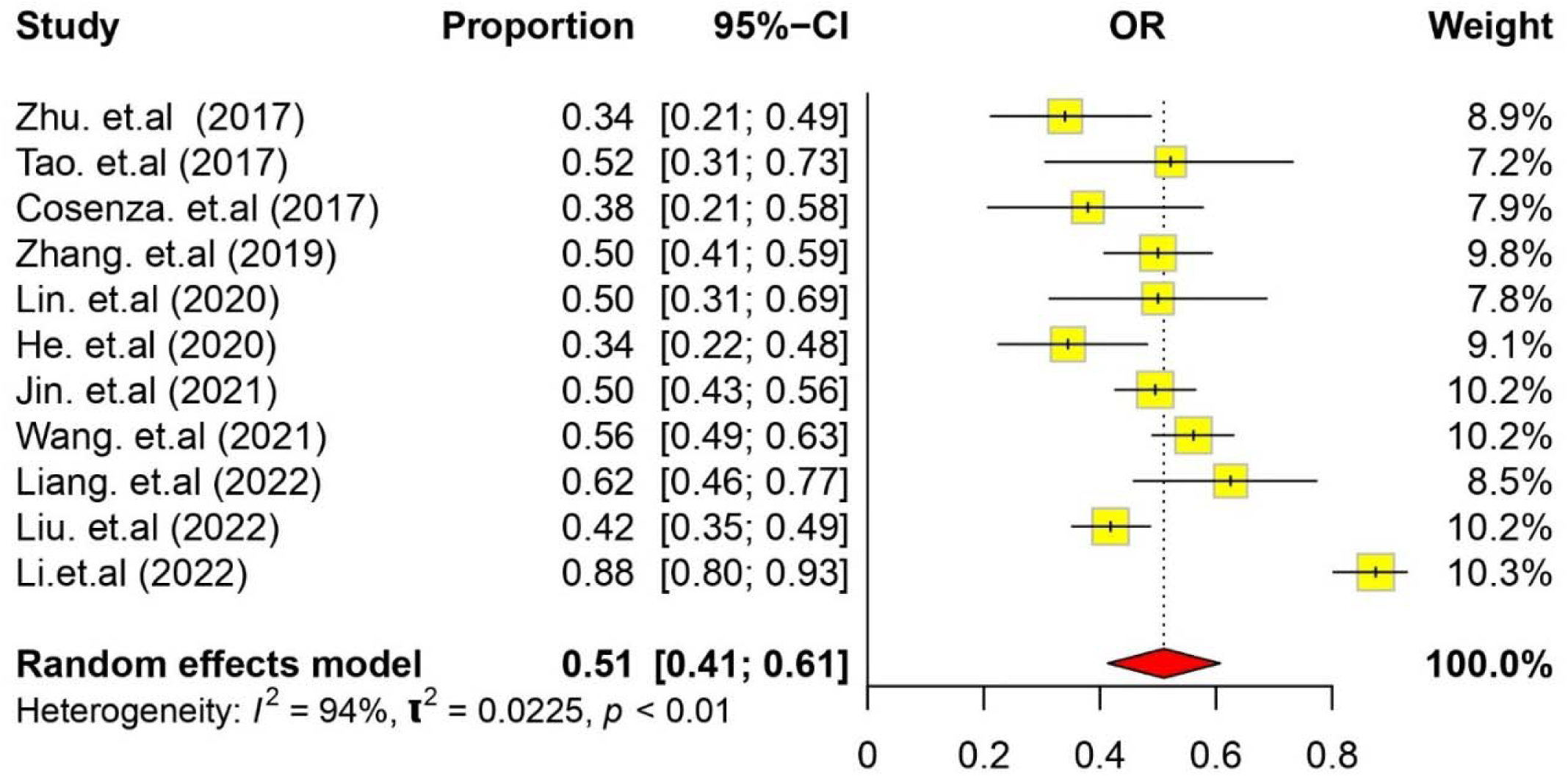
The forest plot illustrates the efficacy rates of human and animal-derived exosomes *in vivo* treatment of OA (with their 95 % confidence intervals).

**Figure 3.**
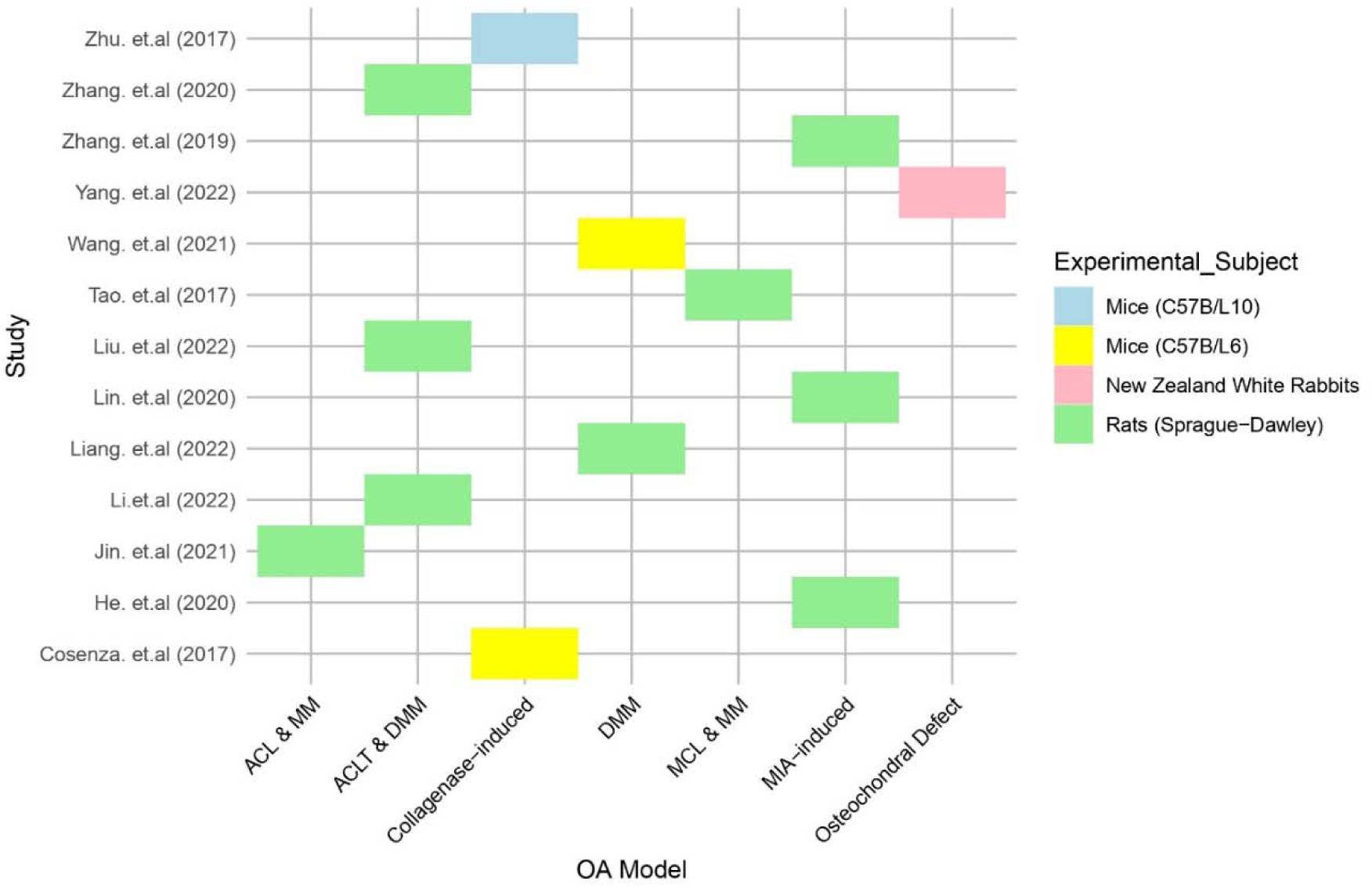
The heatmap illustrates the experimental subjects (on the right side) and osteoarthritis (OA) models (along the x-axis) analyzed across all studies (on the left side of the y-axis).

Both human and animal-derived exosomes demonstrate substantial reductions in OA severity, with human-derived exosomes showing a broader range of ORs from 0.34 (Zhu et al., 2017) to 0.88 (Li et al., 2022), indicating variability but consistently effective outcomes. In contrast, animal-derived exosomes show ORs ranging from 0.34 (He et al., 2020) to 0.62 (Liang et al., 2022), also indicating efficacy but with slightly less variability. Human-derived exosome studies tend to have higher weight contributions (up to 10.3% for Li et al., 2022) compared to animal-derived exosome studies, suggesting a greater overall influence on the meta-analysis outcomes. This overview provides a crucial understanding of the broad impact of exosome-based therapies on OA, offering a snapshot of the therapeutic potential of these treatments across diverse study conditions.

### 3.4. Assessment of Experimental Subjects and Osteoarthritis Models Utilized for Evaluating Exosome Efficacy

A detailed assessment of experimental subjects and osteoarthritis (OA) models was utilized in various studies to evaluate the efficacy of exosome therapy. Each study is represented by a bar that indicates the OA model employed and the type of experimental subject used. The OA models include Anterior Cruciate Ligament Transection (ACL) combined with Medial Meniscectomy (MM), Anterior Cruciate Ligament Transection (ACLT) combined with Destabilization of the Medial Meniscus (DMM), Collagenase-induced OA, DMM alone, Medial Collateral Ligament (MCL) combined with MM, Monosodium Iodoacetate (MIA)-induced OA, and Osteochondral Defect. The experimental subjects are color-coded to distinguish between Mice (C57BL/10), Mice (C57BL/6), New Zealand White Rabbits, and Rats (Sprague-Dawley).

The studies exhibit a diverse range of OA models and experimental subjects. For instance, Zhu et al. (2017) used C57BL/10 mice in an ACL & MM model, while Zhang et al. (2020) utilized C57BL/6 mice in a DMM model. Zhang et al. (2019) and Yang et al. (2022) both employed New Zealand White Rabbits but used different OA models, specifically ACLT & DMM and DMM, respectively. Studies by Wang et al. (2021) and He et al. (2020) used C57BL/6 mice in Collagenase-induced and DMM models, respectively. The use of Sprague-Dawley rats is common in several studies, such as Tao et al. (2017), Liu et al. (2022), Lin et al. (2020), Liang et al. (2022), Li et al. (2022), Jin et al. (2021), and Cosenza et al. (2017). These studies employed a variety of OA models including MIA-induced, Osteochondral Defect, and combinations of ligament transection and meniscectomy. This diversity in experimental design reflects the different approaches to modeling OA and evaluating exosome efficacy, with each model and subject type providing unique insights into the therapeutic potential of exosomes.

Overall, this highlights the diversity in experimental methodologies used across studies assessing exosome therapy for OA. This underlines the importance of considering both the OA model and the experimental subject when interpreting results and comparing outcomes across different research efforts.

### 3.5. Source Analysis of Exosomes from Human and Animal Mesenchymal Stem Cells

The forest plot analysis shown in **Figure 4** provides a more detailed analysis by categorizing the studies based on the source of the exosome’s human versus animal. This subgroup analysis offers a nuanced understanding of how the efficacy of exosome treatments varies depending on their origin. Many of the included publications, whether employing human or animal sources of mesenchymal stem cells, support exosome-based treatment for osteoarthritis. The analysis indicates that exosomes derived from human sources show a slightly higher pooled effect size of 0.53 [0.44; 0.63], compared to 0.44 [0.28; 0.60] for animal-derived exosomes, suggesting that exosomes from human sources may have a marginally greater therapeutic impact on OA.

**Figure 4.**
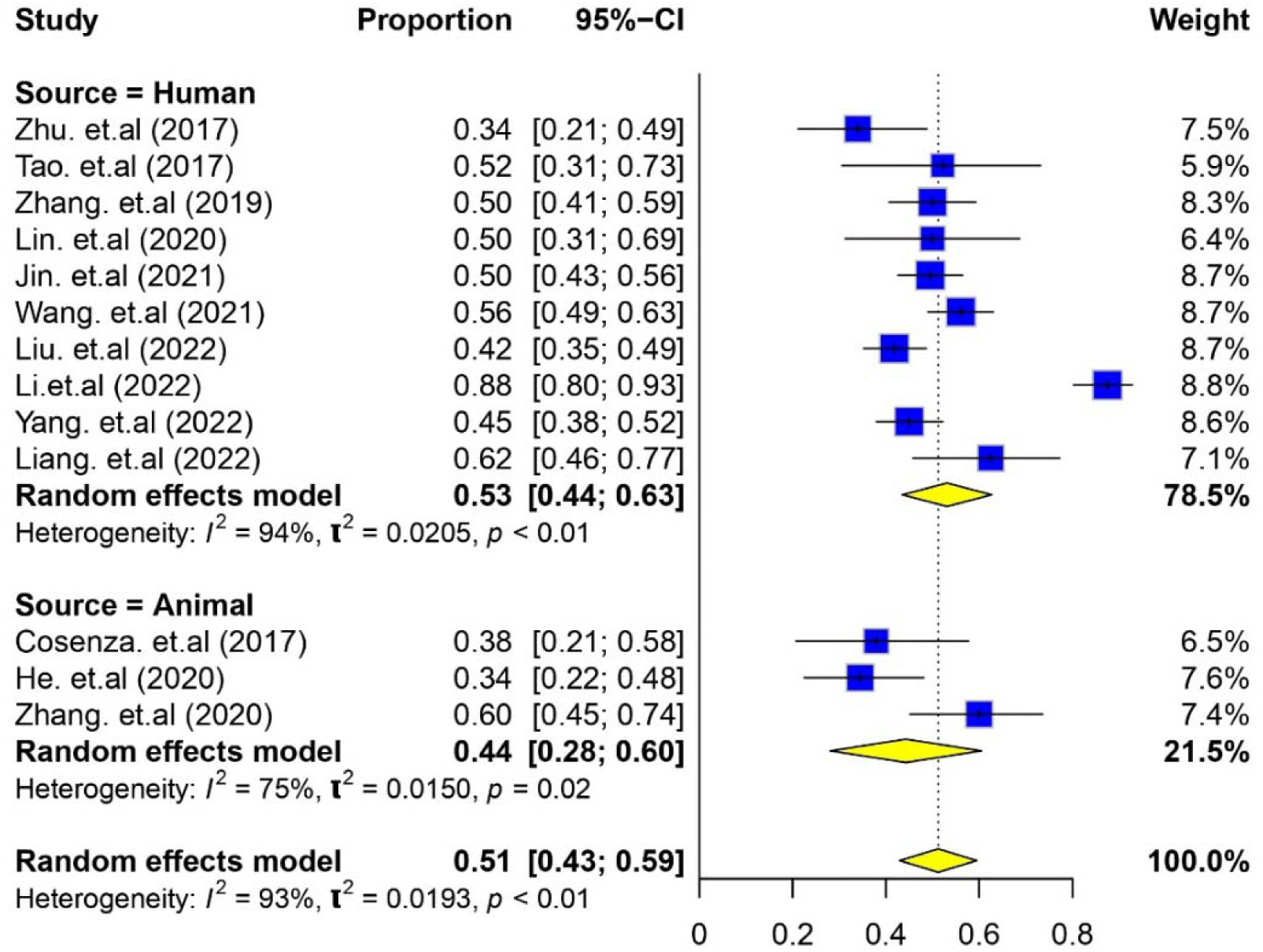
The forest plot displays the efficacy of exosomes derived from human and animal mesenchymal stem cells, as reported in the selected studies.

Moreover, exosomes generated from bone marrow have been particularly noted for their usefulness in the treatment of osteoarthritis, highlighting their potential not only in human applications but also for veterinary purposes. The heterogeneity within each subgroup (I^2^=94% for humans and I^2^=75% for animals) emphasizes the variability in outcomes, with human-derived exosomes showing more inconsistency across studies. This detailed breakdown is essential for clinicians and researchers who are considering the source of exosomes in their therapeutic applications, as it provides insights into the relative efficacy and variability of human versus animal-derived treatments, with bone marrow-derived exosomes showing significant promise.

### 3.6. Risk of bias in the eligible studies

Figure 5 provides a comprehensive risk of bias assessment for 13 included studies using the QUADAS-2 tool (Quality Assessment of Diagnostic Accuracy Studies 2), evaluating five key domains: randomization process (D1), timing of participant recruitment relative to randomization (D1b), deviations from intended intervention (D2), missing outcome data (D3), outcome measurement (D4), and selection of reported results (D5). The majority of studies exhibit a low risk of bias across most domains, particularly in D2, D3, D4, and D5, indicating that these aspects were generally well-controlled. However, several studies (e.g., Wang et al., 2021; Tao et al., 2017) show a high risk of bias in D1, related to the randomization process, raising concerns about internal validity. The D1b domain was not applicable in many cases, reflecting its irrelevance to some study designs. Moderate concerns are noted in a few studies (e.g., Jin et al., 2021) in specific domains, and some studies lack sufficient information to assess certain biases fully, particularly in D1b and D1. Overall, while most studies maintain low to moderate risks of bias, significant issues in the randomization process in some studies highlight the need for rigorous study design and transparent reporting to ensure the reliability of findings in exosome-based osteoarthritis treatments.

**Figure 5.**
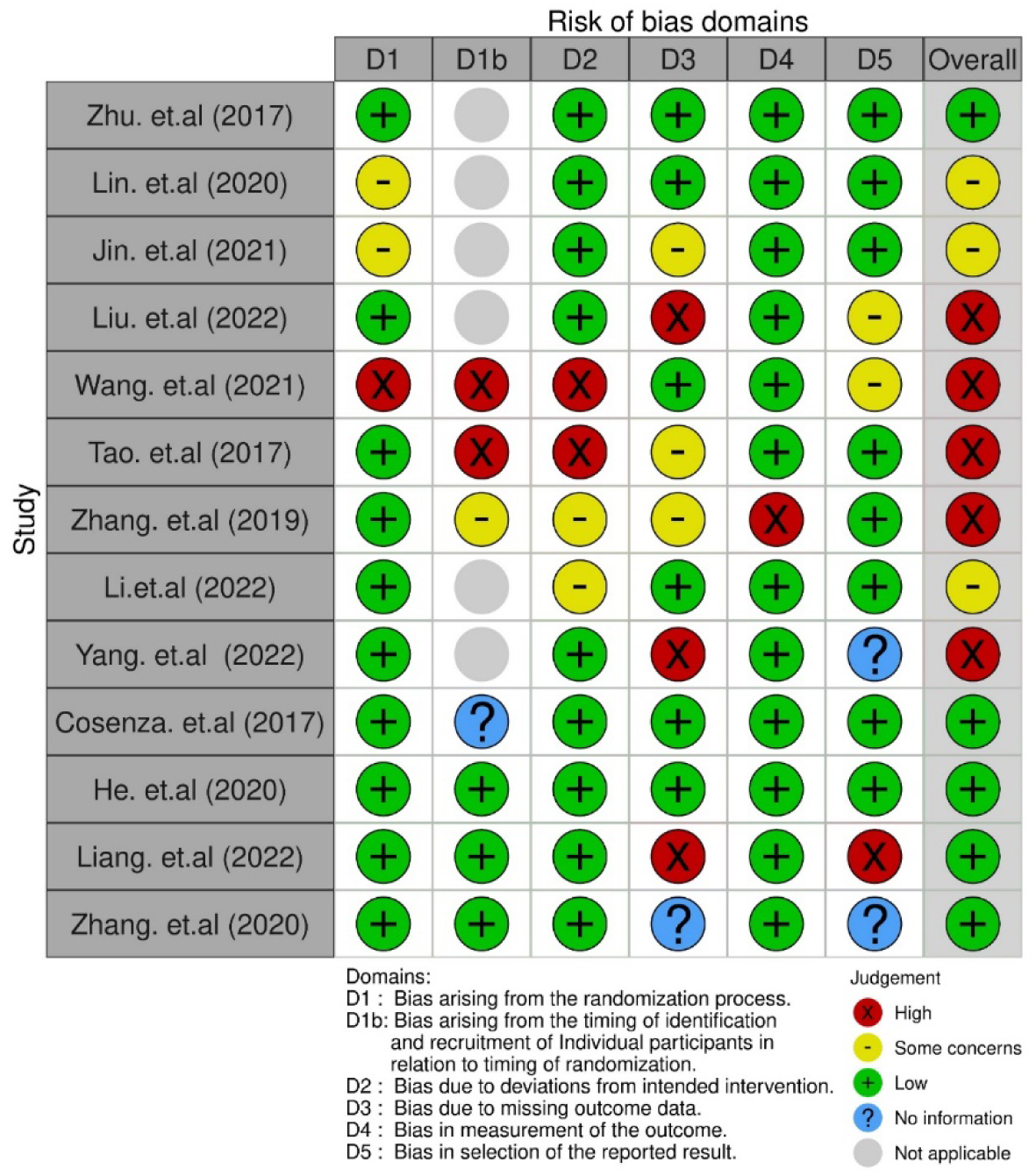
Grouped bar charts illustrating the risk of bias across 13 included studies, were assessed using the QUADAS-2 tool (Quality Assessment of Diagnostic Accuracy Studies-2).

## 4. Discussion

In this study, we rigorously evaluated the efficacy of MSC-derived exosome therapy in mitigating osteoarthritis symptoms in preclinical animal models, while systematically elucidating the mechanisms underlying its therapeutic effects. Our focus on osteoarthritis is justified by its status as the most prevalent degenerative disease, a leading cause of disability, and its strong association with chronic pain and various joint abnormalities, including articular cartilage damage, synovial inflammation, subchondral bone remodeling, and osteophyte formation (Steinmetz et al., 2023; Tong et al., 2022).

We conducted a thorough analysis of 13 peer-reviewed studies published between 2016 and 2023, exploring the therapeutic potential of exosomes derived from both human and animal sources. This analysis was designed to deepen our understanding of exosome therapy’s impact on osteoarthritis progression, accounting for critical factors such as administration methods, dosage frequency, patient age, weight, and OARSI scores. Our findings unequivocally demonstrate several key properties of exosomes, and the experimental models employed. Notably, exosomes, typically ranging in size from 30 to 150 nm, are most effectively administered via the intra-articular route. Despite the clear focus on human osteoarthritis in the literature, a glaring gap persists in research on the application of exosome therapy in veterinary medicine. The reviewed studies consistently show that exosomes significantly reduce the inflammatory response and improve OARSI scores, a crucial measure of osteoarthritis severity encompassing factors like physical function, stiffness, and pain. Remarkably, none of the studies reported adverse effects, underscoring the promise of exosome therapy, particularly as it results in significant reductions in OARSI scores in preclinical settings.

Exosomes play an essential role in modulating immune responses, addressing cardiovascular disorders, combating degenerative diseases, and even influencing cancer progression. Their multifaceted functions, including tissue repair, immune response modulation, and inflammation reduction, position MSC-derived exosomes at the forefront of regenerative medicine (Zakirova et al., 2020). Acting as intercellular messengers, exosomes transport biologically active molecules—such as lipids, proteins, miRNAs, and multimolecular complexes—that are critical in regulating homeostasis and metabolism (Colombo, Raposo, & Théry, 2014). These characteristics make exosomes invaluable for both therapeutic and diagnostic applications, with evidence suggesting that extracellular vesicles alone can effectively heal injuries and repair tissue, thereby circumventing the practical challenges posed by stem cell-based protocols in veterinary medicine (Diomaiuto et al., 2021).

The selection of appropriate animal models is indispensable in osteochondral defect and osteoarthritis research. Our analysis identified the use of various animal models, including mice (C57B/L10, C57B/L6), New Zealand White rabbits, and Sprague-Dawley rats, each offering unique advantages—whether in understanding genetic responses, anatomical similarities to humans, or robustness and availability. The efficacy of exosome therapy for osteoarthritis is evident, irrespective of the MSC source, with exosomes derived from human and animal sources, particularly bone marrow, showing substantial promise in alleviating osteoarthritis symptoms. The forest plot analysis further strengthens the case for exosome-based therapy, with the majority of studies yielding positive results.

Our analysis highlights a critical limitation: the paucity of veterinary models in the existing literature. While research predominantly focuses on human osteoarthritis, there is a conspicuous lack of studies investigating the use of exosomes in veterinary contexts. This gap limits the generalizability of our findings to veterinary health, as the effectiveness of exosome therapy in preclinical animal models may not fully capture the complexities of osteoarthritis in veterinary patients. Additionally, the considerable variability in experimental methodologies, including delivery routes, dosage frequencies, and assessment techniques, introduces potential inconsistencies and complicates the establishment of standardized protocols for exosome therapy in both human and veterinary medicine. The transition from preclinical models to clinical applications presents significant challenges, given the inherent differences between animal models and actual patients, whether human or veterinary, which may limit the direct applicability of the findings.

Given these limitations, future research shifts must focus on exosome therapy in veterinary patients with osteoarthritis. Comparative studies involving a broader range of species—such as dogs, cats, and horses—are essential to fully assess the efficacy and safety of exosome therapy across diverse veterinary populations. Incorporating organ-on-chip (OOC) technology into these investigations is a crucial next step. OOC platforms offer unparalleled insights into the mechanisms and therapeutic benefits of exosome therapy by accurately replicating physiological conditions, enabling real-time monitoring of cellular responses, and facilitating sophisticated disease modeling and high-throughput screening (Kang et al., 2021). This approach has the potential to significantly enhance our understanding of exosome therapy’s impact across different species, thereby improving translational outcomes in veterinary medicine.

Furthermore, the standardization of experimental protocols—such as administration routes, dosage regimens, and outcome measures—is critical for enhancing the comparability and reproducibility of research findings. Establishing consensus guidelines for conducting preclinical exosome therapy studies will strengthen the robustness and credibility of the research, paving the way for more reliable conclusions. As we move from preclinical research to clinical applications, well-designed clinical trials are essential to validate the safety and efficacy of exosome therapy in osteoarthritis treatment for both human and veterinary patients. Such trials will provide definitive evidence of the clinical utility of exosome therapy, laying the groundwork for its broader adoption in medical practice.

## 5. Conclusion

In conclusion, our study unequivocally demonstrates the effectiveness of MSC-derived exosome therapy in preclinical osteoarthritis models, underscoring the significant potential of exosome-based interventions to alleviate osteoarthritis symptoms. Through a rigorous and systematic analysis of existing literature, we have pinpointed critical factors that influence the success of exosome therapy, including the source of mesenchymal stem cells, administration routes, and experimental protocols. While our study acknowledges limitations, such as the underrepresentation of veterinary models and variability in experimental designs, these do not detract from the compelling evidence we present. Exosome therapy emerges as a promising and innovative approach for treating osteoarthritis, poised to transform both human and veterinary medicine. Moving forward, research efforts must intensify to resolve outstanding questions, standardize protocols, and expedite the clinical application of exosome therapy. Our findings contribute decisively to the growing body of evidence supporting the adoption of exosome therapy in osteoarthritis treatment, with far-reaching implications for both public and veterinary health.

## Data Availability

All data produced in the present work are contained in the manuscript

## Ethics approval and consent to participate

’Not applicable’

## Consent for publication

All authors have given their consent to publish

## Availability of data and materials

The authors confirm that the data supporting the findings of this study are available within the article.

## Authors’ contributions

All authors have equally contributed to the conceptualization, methodology, and writing and editing of the manuscript.

## Acknowledgements

This research was funded by VtR Inc-CGU (SCRPD1L0221); DOXABIO-CGU (SCRPD1K0131), and CGU grant (UZRPD1L0011, UZRPD1M0081). We have not used AI in the preparation of the manuscript.

